# Neuroprotective Effects of Vitamin D Supplementation on Outcomes in Traumatic Brain Injury: A Systematic Review and Meta-Analysis

**DOI:** 10.1101/2025.10.07.25337269

**Authors:** Farzan Fahim, Fatemeh Vosoughian, Mahdi Mehmandoost, Hengameh Yousefi, Amirmohammad Bahri, Khatere Mokhtari, Tohid Emami-Meybodi, Iman Sarmadi, Amirhossein Zare, Shahin Naghizadeh, Shideh Moftakhari Hajimirzaei, Sayeh Oveisi, Alireza Zali

## Abstract

**Background:** Traumatic brain injury (TBI) remains a leading cause of morbidity and mortality worldwide, with secondary brain damage driven by inflammation and oxidative stress. Vitamin D is increasingly recognized for potential neuroprotective effects in TBI, while data regarding vitamin E remain limited.

**Objective:** To systematically review and meta-analyze the effects of vitamin D supplementation, and qualitatively review evidence for vitamin E, on clinical and functional outcomes after moderate to severe TBI.

**Methods:** A comprehensive search was carried out in PubMed, Scopus, Embase, Web of Science, and Google Scholar up to January 2025. Studies reporting on vitamin D or E supplementation in clinical TBI were eligible. Risk of bias was assessed using JBI checklists. A meta-analysis of randomized controlled trials reporting pre- and post-treatment GCS scores following vitamin D supplementation was performed with a fixed-effect model.

**Results:** From 4,546 records, nine clinical studies met criteria; three RCTs on vitamin D (n=151 patients) were included in the meta-analysis, which found that vitamin D supplementation significantly improved GCS scores versus controls (SMD□= □1.02, 95% CI: 0.68–1.36, p□< □0.0001; I^2^□= □0%). Narrative analysis suggested that vitamin D may improve functional outcomes, reduce inflammatory biomarkers, and lower mortality in select studies. Evidence for vitamin E in TBI is currently limited to a small number of heterogeneous studies, with early data suggesting possible benefits for acute recovery and oxidative stress reduction, but insufficient for quantitative synthesis.

**Conclusion:** Vitamin D supplementation may confer short-term improvement in neurological and functional outcomes following moderate to severe TBI. Existing evidence for vitamin E is insufficient to support robust conclusions. Larger, rigorously designed RCTs—particularly for vitamin E—are required to clarify effectiveness, optimal dosing, and long-term outcomes.

## Introduction

Traumatic brain injury (TBI) is a major public health concern and remains one of the leading causes of morbidity, mortality, and long-term disability worldwide, affecting nearly 69 million people annually, particularly in low- and middle-income countries (1, 2). The primary mechanical impact is followed by complex secondary injury cascades, characterized by neuroinflammation, oxidative stress, and disruption of blood–brain barrier function—all of which contribute to further neuronal loss and neurological deterioration (3-5). Severity assessment and monitoring of prognosis in TBI are typically performed using the Glasgow Coma Scale (GCS), a widely used tool for both clinical care and research (3).

A growing body of evidence highlights the pivotal roles of inflammatory cytokines and oxidative stress markers—including TNF-α, IL-1β, and IL-6—in determining the severity and outcomes of TBI (6, 7). Despite advances in acute care, options for modulating secondary brain injury and improving patient prognosis remain limited, driving interest in novel therapeutic strategies (8, 9).

Among potential interventions, antioxidant vitamins have gained considerable research attention. Vitamin D, beyond its classical roles in calcium homeostasis, has demonstrated immunomodulatory, anti-inflammatory, and neuroprotective effects in both pre-clinical and clinical studies (10-12). Animal studies indicate that vitamin D supplementation can attenuate cerebral edema, lower oxidative damage, and improve neuronal recovery after TBI (13, 14). Human studies further suggest that vitamin D deficiency is common after TBI, and lower serum vitamin D levels may be associated with increased risk of unfavorable outcomes (15-17). Early phase clinical trials have shown that vitamin D supplementation improves neurological function, reduces inflammatory markers, and may decrease duration of mechanical ventilation and ICU stay in patients with moderate to severe TBI (18-22), although methodological heterogeneity and small sample sizes limit definitive conclusions.

Vitamin E, a fat-soluble antioxidant, is another candidate neuroprotectant investigated mostly in animal models, where it has been shown to reduce lipid peroxidation and improve functional and cognitive outcomes after TBI (15, 23). However, clinical evidence for vitamin E supplementation in TBI remains sparse and heterogeneous, mostly limited to small-scale studies and those employing combination antioxidant regimens (24, 25). A few randomized trials suggest vitamin E may reduce acute oxidative stress and possibly mortality, but the overall quality and consistency of available human data are low (24, 25).

Despite these promising findings, major gaps persist in the literature. Most studies are limited by small sample sizes, lack of standardization in dosing and timing of supplementation, and inadequate reporting of long-term and patient-centered outcomes. Notably, while there is more robust data for vitamin D, evidence for clinical efficacy of vitamin E remains insufficient for meta-analytic synthesis (24, 25).

However, despite encouraging preliminary evidence, robust data from large-scale randomized controlled trials are still lacking. Critical questions remain regarding optimal dosing, timing of supplementation, ideal target populations, long-term functional outcomes, and the comparative efficacy of vitamin D versus vitamin E in TBI. Addressing these important gaps is essential for developing clear, evidence-based clinical recommendations.

Therefore, the present systematic review and meta-analysis aims to (i) quantitatively evaluate the effects of vitamin D supplementation on neurological and clinical outcomes in TBI patients, and (ii) provide a qualitative synthesis of current evidence for vitamin E supplementation. By identifying strengths, limitations, and future research directions, this study seeks to clarify the therapeutic potential of these antioxidants in the management of TBI.

## Methods

### Study Design and Registration

This systematic review and meta-analysis were conducted in accordance with the Preferred Reporting Items for Systematic Reviews and Meta-Analyses (PRISMA) 2020 guidelines. The study protocol was prepared a priori and registered in PROSPERO (Registration ID: 1088575).

### Information Sources and Search Strategy

A comprehensive search was performed in PubMed, Scopus, Embase, Web of Science, and Google Scholar, covering records from database inception until January 31, 2025. The search strategy combined Medical Subject Headings (MeSH) and relevant free-text keywords related to vitamin D, vitamin E, traumatic brain injury (TBI), prognosis, functional recovery, and management. The full search strategy for each database is available in Supplementary Table S1. In addition, the reference lists of all included articles and relevant reviews were screened to identify further eligible studies.

### Eligibility Criteria

#### Studies were included if they

- Enrolled human participants;
- Investigated the effects of vitamin D and/or vitamin E supplementation (including relevant MeSH terms);
- Reported clinical or functional outcomes for adult TBI patients (such as Glasgow Coma Scale [GCS], Glasgow Outcome Scale [GOS/GOS-E], mortality, ICU/hospital stay, or inflammatory/oxidative biomarkers);
- Were published as English-language, full-text original articles.

#### Studies were excluded if they

- Were animal or in vitro investigations, case reports, conference abstracts, reviews, meta-analyses, protocols, or editorials;
- Focused exclusively on other vitamins without separate data for vitamin D or E;
- Lacked extractable or relevant outcome data.

### Study Selection

Duplicate records were removed using EndNote, and the remaining unique articles were imported into Rayyan for title and abstract screening by two independent reviewers. Full texts of potentially eligible studies were then retrieved and assessed according to the inclusion and exclusion criteria. Discrepancies at any stage were resolved by consensus or, if necessary, by consultation with a third reviewer. The study selection process is illustrated in the PRISMA flow diagram (Figure 1).

**Figure 1.**
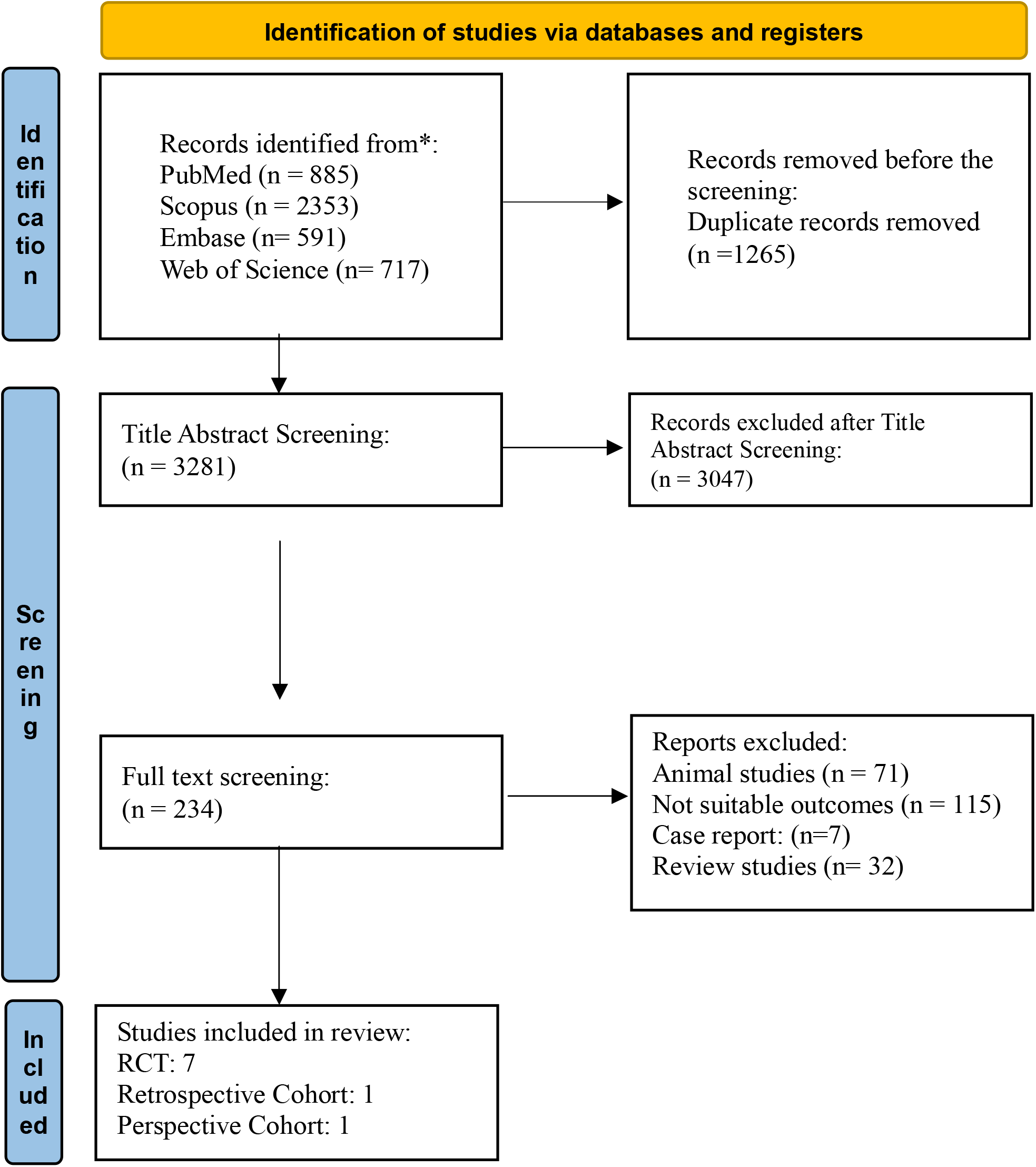
PRISMA Flow chart

**Figure 2.**
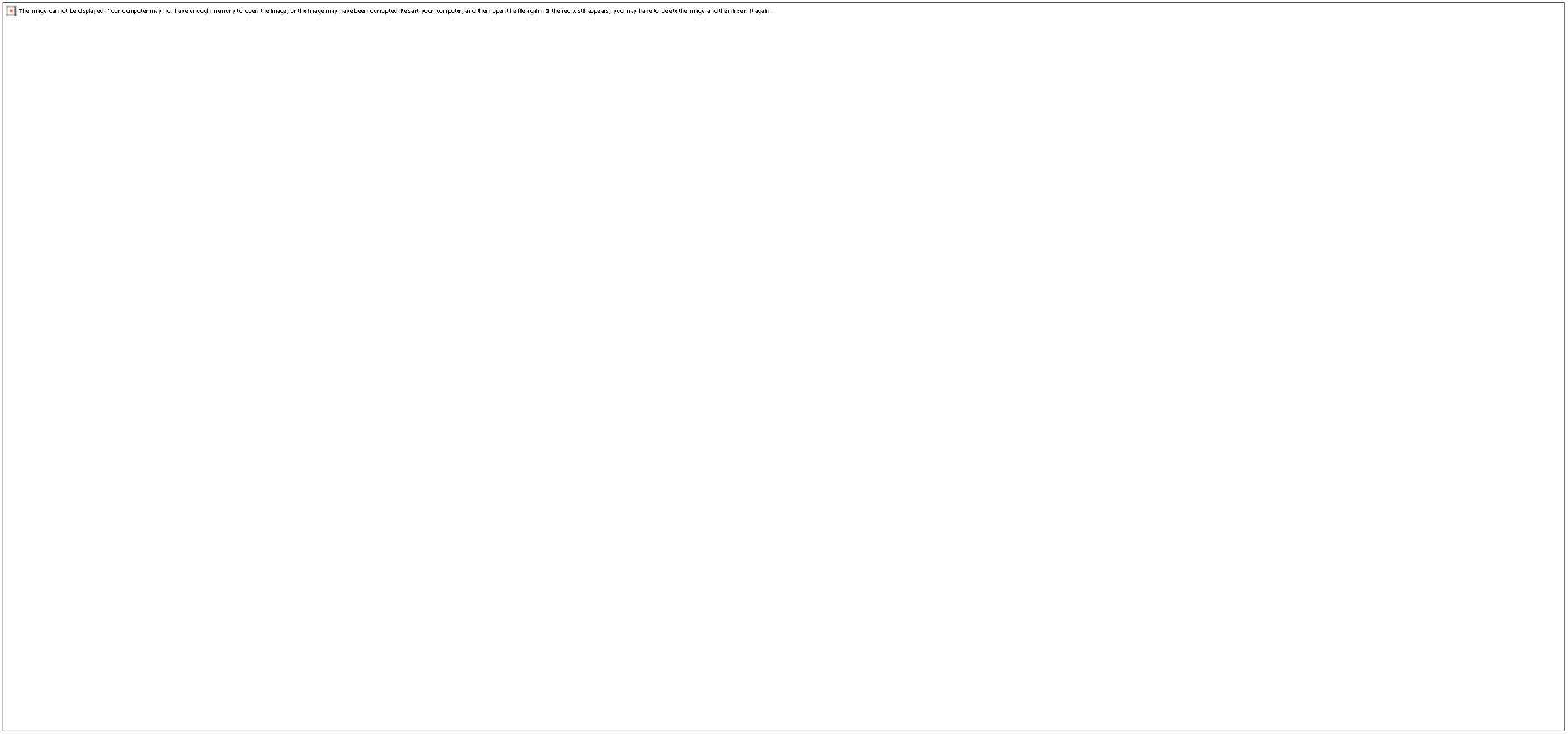
Forest plot showing the standardized mean difference (SMD) in GCS scores between vitamin D-supplemented groups and control groups. A fixed-effect model was used due to low heterogeneity (I^2^ = 0%).

### Data Extraction

Two reviewers independently extracted data using a standardized, pilot-tested data extraction form. The following variables were collected: first author, year, country, study design, sample size, patient demographics (age, sex, TBI severity), intervention details (supplement type, dose, route, duration), comparator(s), relevant clinical outcomes (GCS, GOS/GOS-E, mortality, hospital and ICU stay, mechanical ventilation duration, biomarkers, adverse events), follow-up, funding sources, and reported conflicts of interest. Any discrepancies in data extraction were resolved through discussion or, if necessary, adjudication by a third reviewer.

### Risk of Bias Assessment

Each included study was independently assessed for risk of bias by two reviewers using the appropriate Joanna Briggs Institute (JBI) Critical Appraisal Checklist: the JBI checklist for randomized controlled trials for RCTs and the JBI checklist for cohort studies for cohort designs. Disagreements were resolved through consensus or a third reviewer. No study was excluded based on high risk of bias. The risk of bias assessments are summarized in Table 1.

**Table 1.**
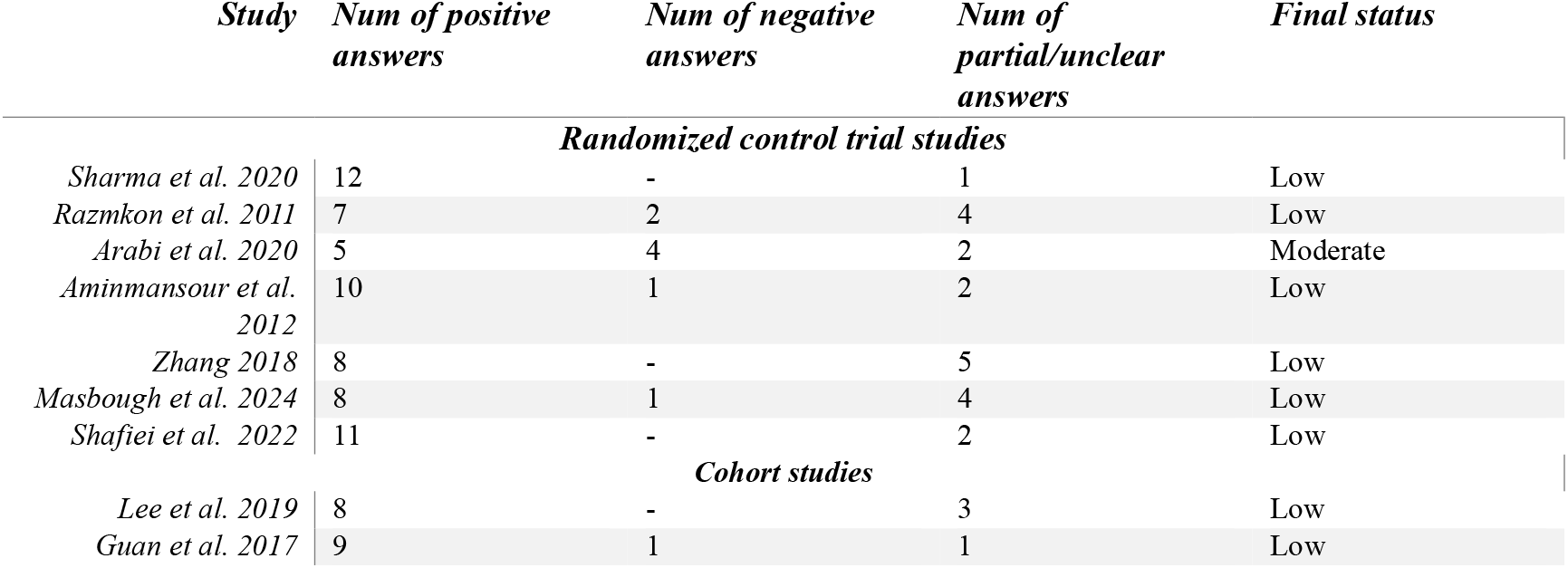

*The risk of bias assessment JBI Critical Appraisal checklists*

### Data Synthesis and Statistical Analysis

A quantitative meta-analysis was performed for randomized controlled trials (RCTs) that reported pre- and post-intervention GCS scores in vitamin D and control groups. Standardized mean differences (SMD) with 95% confidence intervals (CIs) were calculated. Given the low degree of observed heterogeneity (I^2^ < 25%), a fixed-effect model was utilized. Restricted maximum likelihood (REML) estimation and inverse variance weighting were used to compute summary estimates. Heterogeneity was further examined using Cochran’s Q test and the I^2^ statistic.

Robustness of findings was evaluated by leave-one-out sensitivity analyses. Publication bias was assessed through funnel plot inspection and Egger’s regression test. Where data permitted, meta-regression was conducted to assess the influence of age and sex on outcome effect sizes.

All statistical analyses were performed using R (version 4.4.2) with the “meta” and “metafor” packages. For vitamin E supplementation, due to insufficient and heterogeneous evidence, only a qualitative synthesis was conducted.

### Outcomes

#### Primary outcome

- Change in Glasgow Coma Scale (GCS) following vitamin D or E supplementation.

#### Secondary outcomes

- Glasgow Outcome Scale (GOS or GOS-E);
- Mortality;
- Duration of ICU stay, hospital stay, and mechanical ventilation;
- Inflammatory and oxidative stress biomarkers;
- Safety and adverse effects.

### Ethics

No ethics approval was required for this systematic review and meta-analysis as only previously published, de-identified data were used.

### esults

#### Study Selection

A total of 4,546 records were identified through database searching. After removal of 1,265 duplicates, 3,281 unique articles were screened by title and abstract using Rayyan. Of these, 28 articles were assessed in full text, and nine studies were included in the final analysis (Figure 1).

#### Study Characteristics

Nine studies, including six randomized controlled trials and three cohort studies, were published between 2011 and 2024. Sample sizes ranged from 35 to 497 participants. Most studies assessed vitamin D supplementation (oral or intramuscular, with doses ranging from 50,000 to 300,000 IU), while two studies examined vitamin E (intramuscular or intravenous). TBI severity ranged from moderate to severe (admission GCS scores 3–12). Participants were predominantly male (approximately 70–80%), with ages primarily between 30 and 50 years. Full study details are provided in Table 1.

#### Risk of Bias

The Joanna Briggs Institute (JBI) Critical Appraisal checklists showed most studies to be at low risk of bias, with two studies assessed as moderate due to incomplete blinding or outcome reporting (Table 1).

#### Quantitative Synthesis (Meta-Analysis)

Three randomized controlled trials (18, 21, 22) (n=151; intervention: 78, control: 73) reported change in Glasgow Coma Scale (GCS) and were included in the meta-analysis. The pooled standardized mean difference (SMD) for GCS improvement with vitamin D versus control was 1.02 (95% CI: 0.68 to 1.36, p < 0.0001), favoring vitamin D. Heterogeneity was negligible (Q = 1.93, I^2^ = 0%, τ^2^ < 0.0001). Leave-one-out sensitivity analysis confirmed result stability. No publication bias was detected (Egger’s test t = 0.50, p = 0.71; funnel plot in Supplementary Figure S2). Meta-regression found no significant effect of age (estimate = 0.021, p = 0.701) or female proportion (estimate = 16.319, p = 0.178) on treatment outcome **(Table 2, Supplementary Figure S1)**.

**Table 2.**
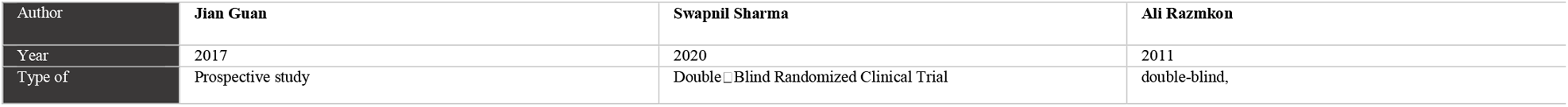
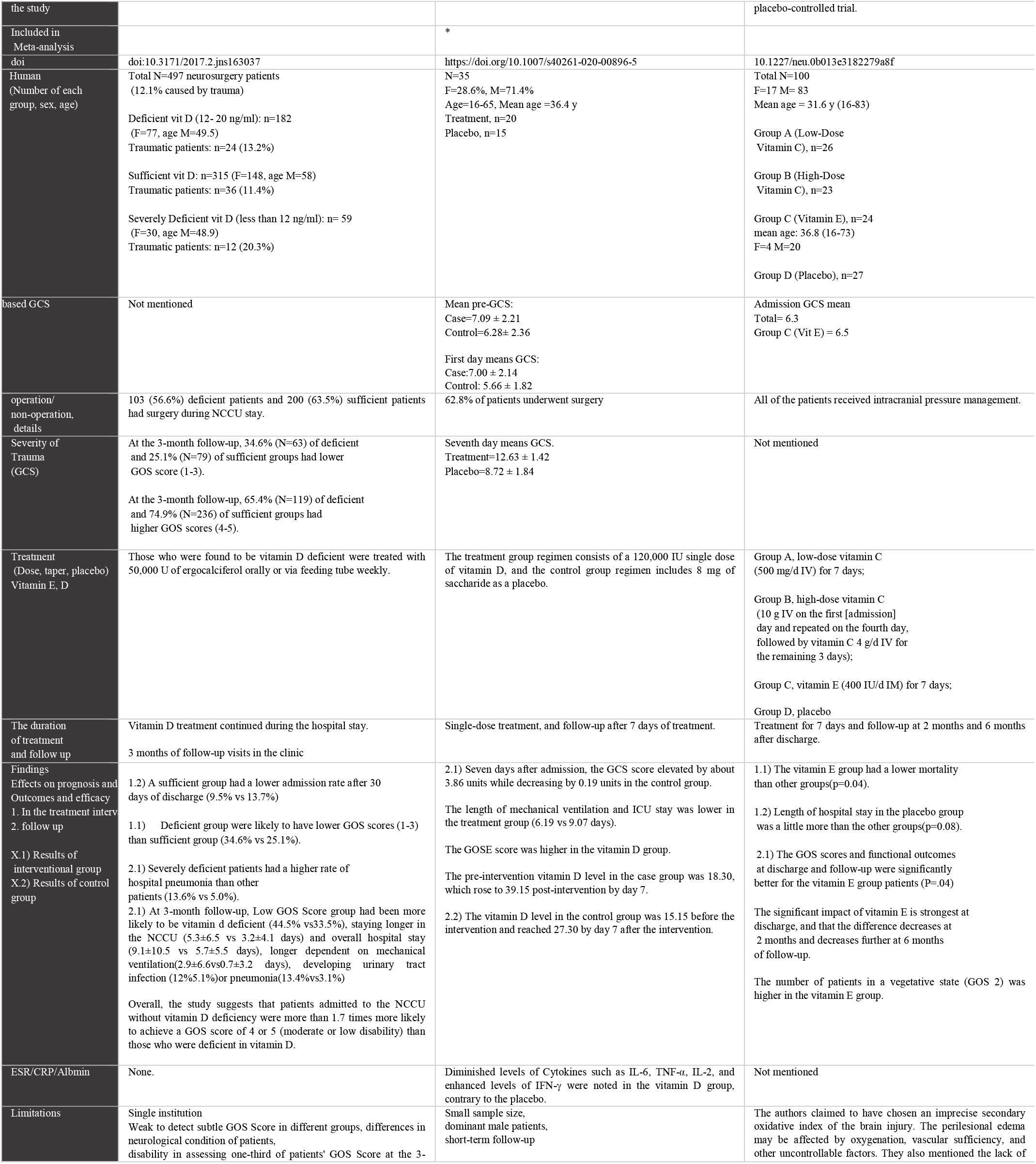
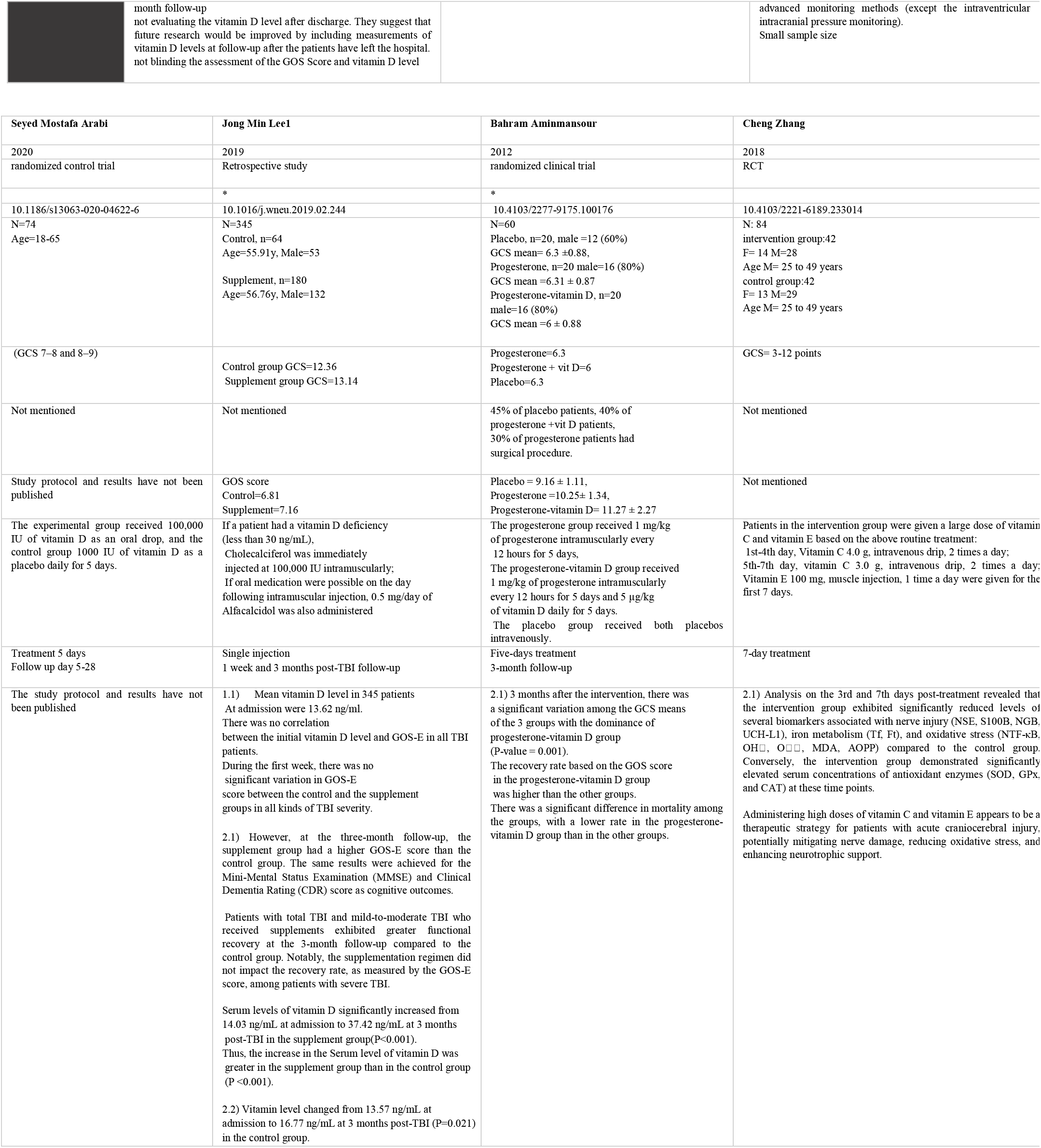
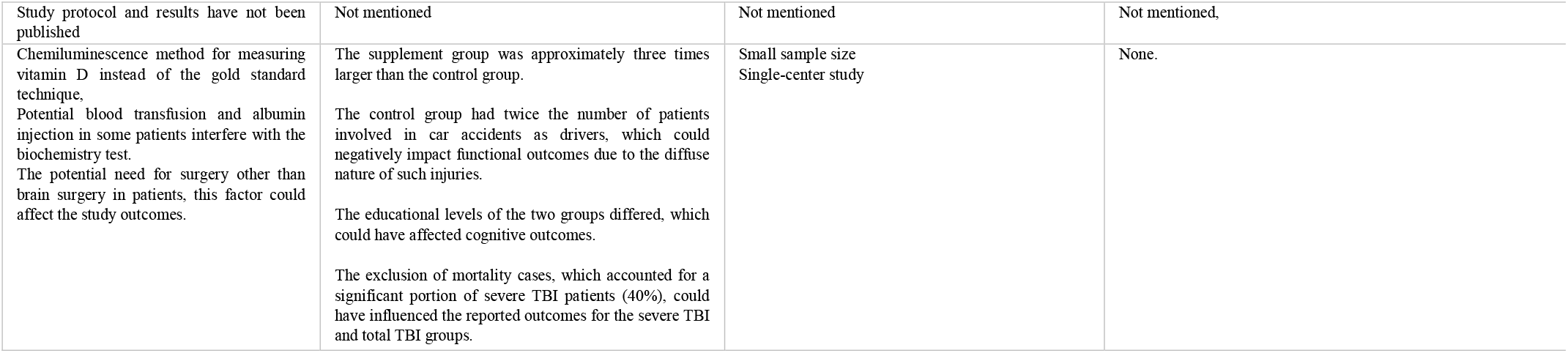
Characteristics of Included Studies.

**Table 3.**
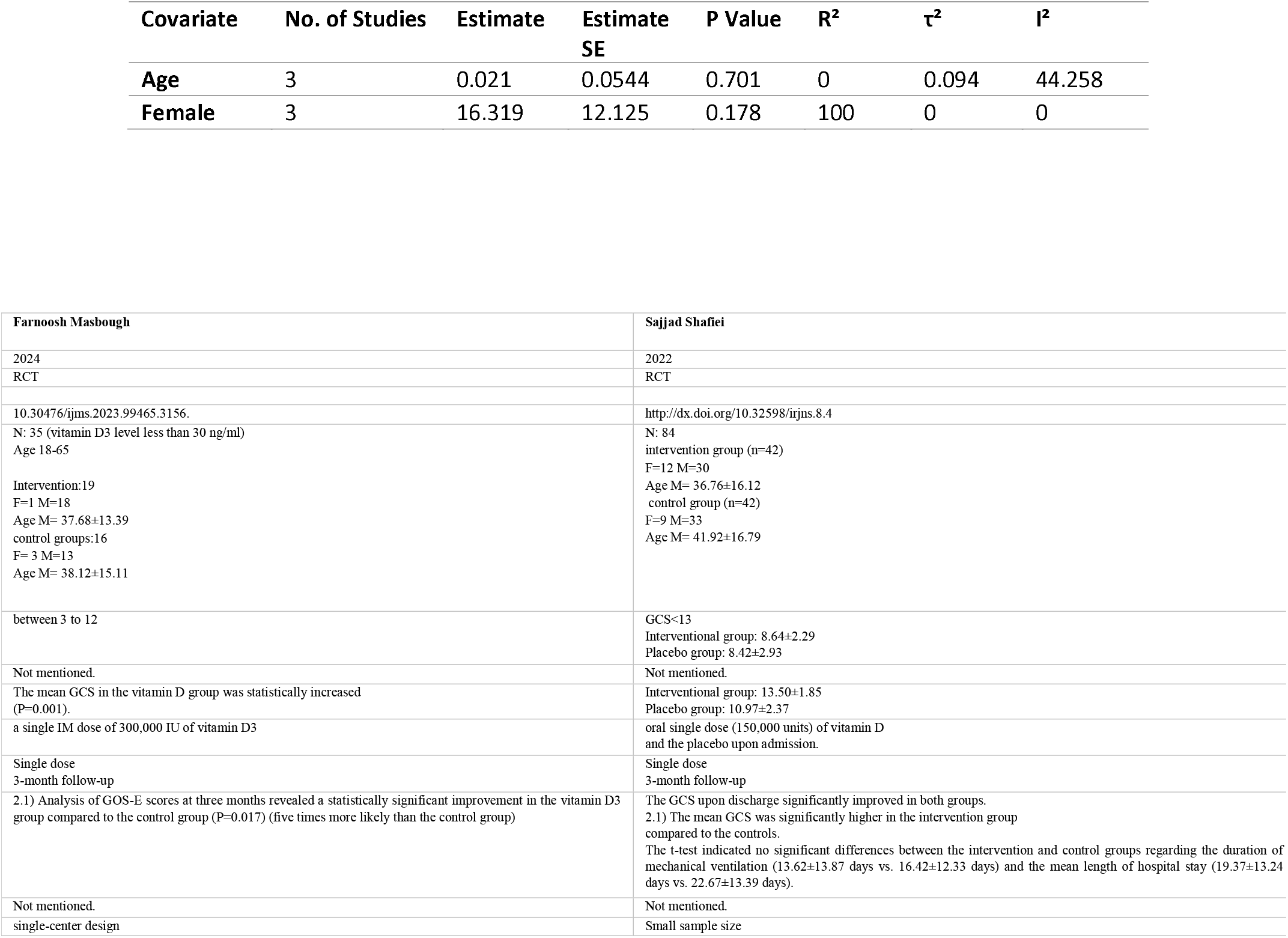
Meta-regression results for the influence of age and gender on GCS outcomes.

| Covariate | No. of Studies | Estimate | Estimate SE | P Value | R<sup>2</sup> | τ<sup>2</sup> | I<sup>2</sup> |
| --- | --- | --- | --- | --- | --- | --- | --- |
| Age | 3 | 0.021 | 0.0544 | 0.701 | 0 | 0.094 | 44.258 |
| Female | 3 | 16.319 | 12.125 | 0.178 | 100 | 0 | 0 |

### Functional and Clinical Outcomes

#### Glasgow Outcome Scale (GOS/GOS-E)

Vitamin D-sufficient patients demonstrated higher rates of favorable GOS at 3 and 6 months post-TBI (17), and Lee et al. (19) observed more patients achieving GOS-E ≥6 in the supplemented group at both timepoints. Masbough et al. (20) found that vitamin D increased odds of favorable GOS-E at three months (P = 0.017). In Razmkon et al. (24), vitamin E led to higher GOS at discharge (P = 0.04); differences faded by later follow-up.

#### Mortality

Lower mortality was observed in the vitamin E group in Razmkon et al. (24) (P = 0.04), and Shafiei et al. (21) reported a lower (but not statistically significant) mortality with vitamin D. Adjunctive vitamin D with progesterone decreased mortality compared to progesterone alone or placebo (18).

### Secondary Outcomes

#### Mechanical Ventilation and ICU/Hospital Stay

Vitamin D significantly reduced mechanical ventilation duration (Sharma et al.: 6.19 ± 1.64 vs. 9.07 ± 2.18 days, P < 0.001 [21]; Guan et al.: 0.7 ± 3.2 vs. 2.9 ± 6.6 days, P = 0.01 (17)). Shafiei et al. (21) and Masbough et al. (20) found no significant difference in ventilation or hospital/ICU stay.

#### Readmission and Complications

Vitamin D-sufficient patients experienced lower 30-day readmission (9.5% vs. 13.7%) and hospital-acquired pneumonia (5.0% vs. 13.6%) (17).

#### Inflammatory and Oxidative Biomarkers

Vitamin D reduced IL-6, TNF-α, and IL-2, and increased IFN-γ in Sharma et al (22). Masbough et al. (20) noted a significant reduction in neutrophil-to-lymphocyte ratio (NLR). Vitamin E (Zhang et al. (25)) reduced markers of oxidative stress (NTF-κB, OH□, O□ □, MDA, AOPP) and nerve injury (NSE, S100B), with increased antioxidant enzymes (SOD, GPx, CAT).

## Discussion

### Summary of Main Findings

In this systematic review and meta-analysis, the efficacy of vitamin D and vitamin E supplementation in patients with traumatic brain injury (TBI) was assessed across nine studies, including six randomized controlled trials and three cohort studies published between 2011 and 2024. Quantitative synthesis of three RCTs demonstrated a significant benefit of vitamin D on neurological recovery, with a pooled SMD of 1.02 (95% CI: 0.68–1.36, p < 0.0001) for GCS improvement, and negligible heterogeneity (I^2^ = 0%). Functional recovery, as measured by the Glasgow Outcome Scale (GOS/GOS-E), also favored vitamin D and E supplementation, with multiple studies reporting higher rates of favorable outcomes and reduced mortality in the intervention groups. Both vitamins showed anti-inflammatory and antioxidant activity, reflected in reduced markers of oxidative stress and inflammatory cytokines.

### Interpretation in the Context of Previous Research

The observed neurological improvement with vitamin D supplementation is consistent with findings from prior clinical and preclinical studies, which have suggested a role for vitamin D in modulating neuroinflammation and promoting neurorecovery (17, 19-22). The anti-inflammatory cytokine profile following vitamin D administration—including reduced IL-6, TNF-α, and IL-2 with increased IFN-γ (20, 22)—may help attenuate the secondary injury cascade, supporting previous reports of its neuroprotective effects (4, 7, 22). Similarly, the positive impact of vitamin D on functional outcomes (as indicated by higher GOS/E scores) aligns with earlier evidence that adequate vitamin D status is associated with better post-TBI prognosis (17, 19).

Vitamin E demonstrated reductions in oxidative stress biomarkers (such as MDA, AOPP, NTF-κB) and neuronal injury markers (NSE, S100B), which concurs with its established antioxidative mechanisms (10, 11, 25). The observed decrease in mortality in vitamin E groups (24) supports the hypothesis that antioxidant therapy may confer survival benefits in severe TBI, as previously reported in related translational studies (11).

Of note, adjunctive treatment with vitamin D and progesterone was associated with lower mortality compared to progesterone alone or placebo (18), suggesting potential synergistic effects, as also noted in experimental models (14, 26). However, not all studies reported significant improvements in all outcomes, and variations in dosing, timing, and study population characteristics likely contributed to heterogeneity.

### Clinical Implications

The present findings highlight the potential role of vitamin D supplementation—and to a lesser extent vitamin E—in improving neurological recovery and reducing complications following moderate-to-severe TBI. Considering the low risk profile and high prevalence of vitamin D deficiency among critically ill patients, routine screening and early correction may be considered as part of neurocritical care protocols (17, 22). Nevertheless, current evidence does not yet support universal high-dose vitamin supplementation for all TBI patients; further individualized assessment remains necessary.

### Limitations

Several limitations must be noted. The total number of high-quality randomized controlled trials remains limited, with only three studies comprising the meta-analysis of GCS outcomes. Sample sizes were small in several studies, reducing statistical power. There was heterogeneity in vitamin dosing regimens (ranging from 50,000 to 300,000 IU for vitamin D), modes of administration (oral, intramuscular, intravenous), and follow-up durations (most limited to three months or less). Some studies combined vitamin supplementation with other interventions (e.g., progesterone), complicating attribution of effects. Risk of bias was generally low, but some studies had issues related to incomplete blinding or outcome reporting. Finally, publication bias cannot be entirely excluded, despite negative findings on formal testing (Egger’s test, funnel plot).

### Future Directions and Conclusion

Larger, multicenter RCTs with standardized vitamin supplementation protocols, longer follow-up, and consistent outcome definitions are required to validate these findings and clarify long-term benefits. Research should also explore the potential for combination therapies (e.g., vitamin D with progesterone) and optimal patient selection.

In summary, evidence supports a beneficial effect of vitamin D—and possibly vitamin E—on early neurological recovery and some clinical outcomes following moderate-to-severe TBI, though routine use awaits confirmation in further high-quality studies.

## Conclusion

This systematic review and meta-analysis provide moderate-quality evidence that vitamin D supplementation, and possibly vitamin E, confer measurable benefits on early neurological recovery and selected clinical outcomes in patients with moderate-to-severe traumatic brain injury. Despite statistically significant improvements—particularly in Glasgow Coma Scale scores—across available randomized trials, the current evidence base is restricted by methodological heterogeneity, limited sample sizes, and predominantly short-term follow-up. Accordingly, while routine correction of vitamin D deficiency may be justified as part of comprehensive neurocritical care, universal high-dose supplementation cannot yet be broadly recommended. Further large-scale, rigorously designed clinical trials are warranted to clarify optimal dosing strategies, long-term efficacy, and potential synergistic effects with other neuroprotective agents.

## Data Availability

All data produced in the present work are contained in the manuscript

## Declarations

### Ethical approval and consent to participate

Not applicable

### Consent for publication

All authors approve this statement

### Availability of data and materials

Not applicable

## Author contributions

Conceptualization: F.V, Data curation: H.Y, M.M, Formal analysis: M.M, A.Z, Investigation: I.S, T.E, K.M Methodology: K.M, A.B, Project administration: F.F, Writing – original draft: M.M, Sh.M, Sh.N, S.O, Writing – review and editing: F.V, A.Z.

## Acknowledgements

Not applicable

## Preprint

https://doi.org/10.1101/2025.10.07.25337269

