## Supplementory for "Neuroprotective Effects of Vitamin D Supplementation on Outcomes in Traumatic Brain Injury: A Systematic Review and Meta-Analysis"

| uthor | **Jian Guan** | **Swapnil Sharma** | **Ali Razmkon** |
| --- | --- | --- | --- |
| Year | 2017 | 2020 | 2011 |
| Type of  the study | Prospective study | Double‑Blind Randomized Clinical Trial | double-blind,  placebo-controlled trial. |
| Included in  Meta-analysis |  | * |  |
| doi | doi:10.3171/2017.2.jns163037 | https://doi.org/10.1007/s40261-020-00896-5 | 10.1227/neu.0b013e3182279a8f |
| Human  (Number of each  group, sex, age) | Total N=497 neurosurgery patients  (12.1% caused by trauma)  Deficient vit D (12- 20 ng/ml): n=182  (F=77, age M=49.5)  Traumatic patients: n=24 (13.2%)  Sufficient vit D: n=315 (F=148, age M=58)  Traumatic patients: n=36 (11.4%)  Severely Deficient vit D (less than 12 ng/ml): n= 59  (F=30, age M=48.9)  Traumatic patients: n=12 (20.3%) | N=35  F=28.6%, M=71.4%  Age=16-65, Mean age =36.4 y  Treatment, n=20  Placebo, n=15 | Total N=100  F=17 M= 83  Mean age = 31.6 y (16-83)  Group A (Low-Dose  Vitamin C), n=26  Group B (High-Dose  Vitamin C), n=23  Group C (Vitamin E), n=24  mean age: 36.8 (16-73)  F=4 M=20  Group D (Placebo), n=27 |
| based GCS | Not mentioned | Mean pre-GCS:  Case=7.09 ± 2.21  Control=6.28± 2.36  First day means GCS:  Case:7.00 ± 2.14  Control: 5.66 ± 1.82 | Admission GCS mean  Total= 6.3  Group C (Vit E) = 6.5 |
| operation/  non-operation,  details | 103 (56.6%) deficient patients and 200 (63.5%) sufficient patients had surgery during NCCU stay. | 62.8% of patients underwent surgery | All of the patients received intracranial pressure management. |
| Severity of  Trauma  (GCS) | At the 3-month follow-up, 34.6% (N=63) of deficient  and 25.1% (N=79) of sufficient groups had lower  GOS score (1-3).  At the 3-month follow-up, 65.4% (N=119) of deficient  and 74.9% (N=236) of sufficient groups had  higher GOS scores (4-5). | Seventh day means GCS.  Treatment=12.63 ± 1.42  Placebo=8.72 ± 1.84 | Not mentioned |
| Treatment  (Dose, taper, placebo)  Vitamin E, D | Those who were found to be vitamin D deficient were treated with 50,000 U of ergocalciferol orally or via feeding tube weekly. | The treatment group regimen consists of a 120,000 IU single dose of vitamin D, and the control group regimen includes 8 mg of saccharide as a placebo. | Group A, low-dose vitamin C  (500 mg/d IV) for 7 days;  Group B, high-dose vitamin C  (10 g IV on the first [admission]  day and repeated on the fourth day,  followed by vitamin C 4 g/d IV for  the remaining 3 days);  Group C, vitamin E (400 IU/d IM) for 7 days;  Group D, placebo |
| The duration  of treatment  and follow up | Vitamin D treatment continued during the hospital stay.  3 months of follow-up visits in the clinic | Single-dose treatment, and follow-up after 7 days of treatment. | Treatment for 7 days and follow-up at 2 months and 6 months after discharge. |
| Findings  Effects on prognosis and  Outcomes and efficacy  1. In the treatment interval  2. follow up  X.1) Results of  interventional group  X.2) Results of control  group | 1.2) A sufficient group had a lower admission rate after 30  days of discharge (9.5% vs 13.7%)   1.1) Deficient group were likely to have lower GOS scores (1-3)   than sufficient group (34.6% vs 25.1%).  2.1) Severely deficient patients had a higher rate of  hospital pneumonia than other  patients (13.6% vs 5.0%).  2.1) At 3-month follow-up, Low GOS Score group had been more likely to be vitamin d deficient (44.5% vs33.5%), staying longer in the NCCU (5.3±6.5 vs 3.2±4.1 days) and overall hospital stay (9.1±10.5 vs 5.7±5.5 days), longer dependent on mechanical ventilation(2.9±6.6vs0.7±3.2 days), developing urinary tract infection (12%5.1%)or pneumonia(13.4%vs3.1%)  Overall, the study suggests that patients admitted to the NCCU without vitamin D deficiency were more than 1.7 times more likely to achieve a GOS score of 4 or 5 (moderate or low disability) than those who were deficient in vitamin D. | 2.1) Seven days after admission, the GCS score elevated by about 3.86 units while decreasing by 0.19 units in the control group.  The length of mechanical ventilation and ICU stay was lower in the treatment group (6.19 vs 9.07 days).  The GOSE score was higher in the vitamin D group.  The pre-intervention vitamin D level in the case group was 18.30, which rose to 39.15 post-intervention by day 7.  2.2) The vitamin D level in the control group was 15.15 before the intervention and reached 27.30 by day 7 after the intervention. | 1.1) The vitamin E group had a lower mortality  than other groups(p=0.04).  1.2) Length of hospital stay in the placebo group  was a little more than the other groups(p=0.08).  2.1) The GOS scores and functional outcomes  at discharge and follow-up were significantly  better for the vitamin E group patients (P=.04)  The significant impact of vitamin E is strongest at  discharge, and that the difference decreases at  2 months and decreases further at 6 months  of follow-up.  The number of patients in a vegetative state (GOS 2) was higher in the vitamin E group. |
| ESR/CRP/Albmin | None. | Diminished levels of Cytokines such as IL-6, TNF-α, IL-2, and enhanced levels of IFN-γ were noted in the vitamin D group, contrary to the placebo. | Not mentioned |
| Limitations | Single institution  Weak to detect subtle GOS Score in different groups, differences in neurological condition of patients,  disability in assessing one-third of patients' GOS Score at the 3-month follow-up  not evaluating the vitamin D level after discharge. They suggest that future research would be improved by including measurements of vitamin D levels at follow-up after the patients have left the hospital.  not blinding the assessment of the GOS Score and vitamin D level | Small sample size,  dominant male patients,  short-term follow-up | The authors claimed to have chosen an imprecise secondary oxidative index of the brain injury. The perilesional edema may be affected by oxygenation, vascular sufficiency, and other uncontrollable factors. They also mentioned the lack of advanced monitoring methods (except the intraventricular intracranial pressure monitoring).  Small sample size |

| **Seyed Mostafa Arabi** | **Jong Min Lee1** | **Bahram Aminmansour** | **Cheng Zhang** |
| --- | --- | --- | --- |
| 2020 | 2019 | 2012 | 2018 |
| randomized control trial | Retrospective study | randomized clinical trial | RCT |
|  | * | * |  |
| 10.1186/s13063-020-04622-6 | 10.1016/j.wneu.2019.02.244 | 10.4103/2277-9175.100176 | 10.4103/2221-6189.233014 |
| N=74  Age=18-65 | N=345  Control, n=64  Age=55.91y, Male=53  Supplement, n=180  Age=56.76y, Male=132 | N=60  Placebo, n=20, male =12 (60%)  GCS mean= 6.3 ±0.88,  Progesterone, n=20 male=16 (80%)  GCS mean =6.31 ± 0.87  Progesterone-vitamin D, n=20  male=16 (80%)  GCS mean =6 ± 0.88 | N: 84  intervention group:42  F= 14 M=28  Age M= 25 to 49 years  control group:42  F= 13 M=29  Age M= 25 to 49 years |
| (GCS 7–8 and 8–9) | Control group GCS=12.36  Supplement group GCS=13.14 | Progesterone=6.3  Progesterone + vit D=6  Placebo=6.3 | GCS= 3-12 points |
| Not mentioned | Not mentioned | 45% of placebo patients, 40% of  progesterone +vit D patients,  30% of progesterone patients had  surgical procedure. | Not mentioned |
| Study protocol and results have not been published | GOS score  Control=6.81  Supplement=7.16 | Placebo = 9.16 ± 1.11,  Progesterone =10.25± 1.34,  Progesterone-vitamin D= 11.27 ± 2.27 | Not mentioned |
| The experimental group received 100,000 IU of vitamin D as an oral drop, and the control group 1000 IU of vitamin D as a placebo daily for 5 days. | If a patient had a vitamin D deficiency  (less than 30 ng/mL),  Cholecalciferol was immediately  injected at 100,000 IU intramuscularly;  If oral medication were possible on the day  following intramuscular injection, 0.5 mg/day of  Alfacalcidol was also administered | The progesterone group received 1 mg/kg  of progesterone intramuscularly every  12 hours for 5 days,  The progesterone-vitamin D group received  1 mg/kg of progesterone intramuscularly  every 12 hours for 5 days and 5 µg/kg  of vitamin D daily for 5 days.  The placebo group received both placebos intravenously. | Patients in the intervention group were given a large dose of vitamin C and vitamin E based on the above routine treatment:  1st-4th day, Vitamin C 4.0 g, intravenous drip, 2 times a day;  5th-7th day, vitamin C 3.0 g, intravenous drip, 2 times a day; Vitamin E 100 mg, muscle injection, 1 time a day were given for the first 7 days. |
| Treatment 5 days  Follow up day 5-28 | Single injection  1 week and 3 months post-TBI follow-up | Five-days treatment  3-month follow-up | 7-day treatment |
| The study protocol and results have not been published | 1.1) Mean vitamin D level in 345 patients   At admission were 13.62 ng/ml.  There was no correlation  between the initial vitamin D level and GOS-E in all TBI  patients.  During the first week, there was no  significant variation in GOS-E  score between the control and the supplement  groups in all kinds of TBI severity.  2.1) However, at the three-month follow-up, the supplement group had a higher GOS-E score than the control group. The same results were achieved for the Mini-Mental Status Examination (MMSE) and Clinical Dementia Rating (CDR) score as cognitive outcomes.  Patients with total TBI and mild-to-moderate TBI who received supplements exhibited greater functional recovery at the 3-month follow-up compared to the control group. Notably, the supplementation regimen did not impact the recovery rate, as measured by the GOS-E score, among patients with severe TBI.  Serum levels of vitamin D significantly increased from  14.03 ng/mL at admission to 37.42 ng/mL at 3 months  post-TBI in the supplement group(P<0.001).  Thus, the increase in the Serum level of vitamin D was  greater in the supplement group than in the control group  (P <0.001).  2.2) Vitamin level changed from 13.57 ng/mL at  admission to 16.77 ng/mL at 3 months post-TBI (P=0.021)  in the control group. | 2.1) 3 months after the intervention, there was  a significant variation among the GCS means  of the 3 groups with the dominance of  progesterone-vitamin D group  (P-value = 0.001).  The recovery rate based on the GOS score  in the progesterone-vitamin D group  was higher than the other groups.  There was a significant difference in mortality among the groups, with a lower rate in the progesterone-vitamin D group than in the other groups. | 2.1) Analysis on the 3rd and 7th days post-treatment revealed that the intervention group exhibited significantly reduced levels of several biomarkers associated with nerve injury (NSE, S100B, NGB, UCH-L1), iron metabolism (Tf, Ft), and oxidative stress (NTF-κB, OH⁻, O₂⁻, MDA, AOPP) compared to the control group. Conversely, the intervention group demonstrated significantly elevated serum concentrations of antioxidant enzymes (SOD, GPx, and CAT) at these time points.  Administering high doses of vitamin C and vitamin E appears to be a therapeutic strategy for patients with acute craniocerebral injury, potentially mitigating nerve damage, reducing oxidative stress, and enhancing neurotrophic support. |
| Study protocol and results have not been published | Not mentioned | Not mentioned | Not mentioned, |
| Chemiluminescence method for measuring vitamin D instead of the gold standard technique,  Potential blood transfusion and albumin injection in some patients interfere with the biochemistry test.  The potential need for surgery other than brain surgery in patients, this factor could affect the study outcomes. | The supplement group was approximately three times larger than the control group.  The control group had twice the number of patients involved in car accidents as drivers, which could negatively impact functional outcomes due to the diffuse nature of such injuries.  The educational levels of the two groups differed, which could have affected cognitive outcomes.  The exclusion of mortality cases, which accounted for a significant portion of severe TBI patients (40%), could have influenced the reported outcomes for the severe TBI and total TBI groups. | Small sample size  Single-center study | None. |

| **Farnoosh Masbough** | **Sajjad Shafiei** |
| --- | --- |
| 2024 | 2022 |
| RCT | RCT |
| 10.30476/ijms.2023.99465.3156. | http://dx.doi.org/10.32598/irjns.8.4 |
| N: 35 (vitamin D3 level less than 30 ng/ml)  Age 18-65  Intervention:19  F=1 M=18  Age M= 37.68±13.39  control groups:16  F= 3 M=13  Age M= 38.12±15.11 | N: 84  intervention group (n=42)  F=12 M=30  Age M= 36.76±16.12  control group (n=42)  F=9 M=33  Age M= 41.92±16.79 |
| between 3 to 12 | GCS<13  Interventional group: 8.64±2.29  Placebo group: 8.42±2.93 |
| Not mentioned. | Not mentioned. |
| The mean GCS in the vitamin D group was statistically increased  (P=0.001). | Interventional group: 13.50±1.85  Placebo group: 10.97±2.37 |
| a single IM dose of 300,000 IU of vitamin D3 | oral single dose (150,000 units) of vitamin D  and the placebo upon admission. |
| Single dose  3-month follow-up | Single dose  3-month follow-up |
| 2.1) Analysis of GOS-E scores at three months revealed a statistically significant improvement in the vitamin D3 group compared to the control group (P=0.017) (five times more likely than the control group) | The GCS upon discharge significantly improved in both groups.  2.1) The mean GCS was significantly higher in the intervention group  compared to the controls.  The t-test indicated no significant differences between the intervention and control groups regarding the duration of mechanical ventilation (13.62±13.87 days vs. 16.42±12.33 days) and the mean length of hospital stay (19.37±13.24 days vs. 22.67±13.39 days). |
| Not mentioned. | Not mentioned. |
| single-center design | Small sample size |
