## Supplementory for "Neuroprotective Effects of Vitamin D Supplementation on Outcomes in Traumatic Brain Injury: A Systematic Review and Meta-Analysis"

**Identification of studies via databases and registers**

**Screening**

Records identified from*:

PubMed (n = 885)

Scopus (n = 2353)

Embase (n= 591)

Web of Science (n= 717)

Records removed before the screening:

Duplicate records removed (n =1265)

Title Abstract Screening:

(n = 3281)

Records excluded after Title Abstract Screening:

(n = 3047)

Full text screening:

(n = 234)

Studies included in review:

RCT: 7

Retrospective Cohort: 1

Perspective Cohort: 1

**Included**

**Identification**

Reports excluded:

Animal studies (n = 71)

Not suitable outcomes (n = 115)

Case report: (n=7)

Review studies (n= 32)

***Figure 1.*** *PRISMA Flow chart*

***Figure 2****. Forest plot showing the standardized mean difference (SMD) in GCS scores between vitamin D-supplemented groups and control groups. A fixed-effect model was used due to low heterogeneity (I² = 0%).*
