## Supplementory for "Neuroprotective Effects of Vitamin D Supplementation on Outcomes in Traumatic Brain Injury: A Systematic Review and Meta-Analysis"

| **Section and Topic** | **Item #** | **Checklist item** | **Location where item is reported** |
| --- | --- | --- | --- |
| **TITLE** | | |  |
| Title | 1 | Identify the report as a systematic review. | Title page 'Neuroprotective Effects of Vitamin D Supplementation on Outcomes in Traumatic Brain Injury: A Systematic Review and Meta-Analysis' |
| **ABSTRACT** | | |  |
| Abstract | 2 | See the PRISMA 2020 for Abstracts checklist. | Abstract section fully reported (Background, Objective, Methods, Results, Conclusion). |
| **INTRODUCTION** | | |  |
| Rationale | 3 | Describe the rationale for the review in the context of existing knowledge. | Introduction paragraphs 1–3, rationale for the review in the context of existing knowledge. |
| Objectives | 4 | Provide an explicit statement of the objective(s) or question(s) the review addresses. | End of Introduction final paragraph explicitly states objectives (vitamin D meta-analysis and vitamin E qualitative review). |
| **METHODS** | | |  |
| Eligibility criteria | 5 | Specify the inclusion and exclusion criteria for the review and how studies were grouped for the syntheses. | Methods 'Eligibility Criteria' subsection. |
| Information sources | 6 | Specify all databases, registers, websites, organisations, reference lists and other sources searched or consulted to identify studies. Specify the date when each source was last searched or consulted. | Methods 'Information Sources and Search Strategy' subsection (PubMed, Scopus, Embase, Web of Science, Google Scholar; up to Jan 2025). |
| Search strategy | 7 | Present the full search strategies for all databases, registers and websites, including any filters and limits used. | Methods same subsection; full strategies in Supplementary Table S1. |
| Selection process | 8 | Specify the methods used to decide whether a study met the inclusion criteria of the review, including how many reviewers screened each record and each report retrieved, whether they worked independently, and if applicable, details of automation tools used in the process. | Methods 'Study Selection' subsection; two independent reviewers using Rayyan. |
| Data collection process | 9 | Specify the methods used to collect data from reports, including how many reviewers collected data from each report, whether they worked independently, any processes for obtaining or confirming data from study investigators, and if applicable, details of automation tools used in the process. | Methods 'Data Extraction' subsection; two reviewers independently extracted data. |
| Data items | 10a | List and define all outcomes for which data were sought. Specify whether all results that were compatible with each outcome domain in each study were sought (e.g. for all measures, time points, analyses), and if not, the methods used to decide which results to collect. | Methods 'Outcomes' subsection; primary outcome: GCS; secondary: GOS, mortality, biomarkers, etc. |
|  | 10b | List and define all other variables for which data were sought (e.g. participant and intervention characteristics, funding sources). Describe any assumptions made about any missing or unclear information. | Methods 'Data Extraction' subsection; collected study design, demographics, interventions, funding, conflicts, etc. |
| Study risk of bias assessment | 11 | Specify the methods used to assess risk of bias in the included studies, including details of the tool(s) used, how many reviewers assessed each study and whether they worked independently, and if applicable, details of automation tools used in the process. | Methods 'Risk of Bias Assessment' subsection; JBI checklist for RCTs and cohorts. |
| Effect measures | 12 | Specify for each outcome the effect measure(s) (e.g. risk ratio, mean difference) used in the synthesis or presentation of results. | Methods 'Data Synthesis and Statistical Analysis'; standardized mean difference (SMD) used. |
| Synthesis methods | 13a | Describe the processes used to decide which studies were eligible for each synthesis (e.g. tabulating the study intervention characteristics and comparing against the planned groups for each synthesis (item #5)). | Methods 'Data Synthesis and Statistical Analysis' and 'Eligibility Criteria'. |
|  | 13b | Describe any methods required to prepare the data for presentation or synthesis, such as handling of missing summary statistics, or data conversions. | Methods 'Data Synthesis and Statistical Analysis'; describes data handling and conversion. |
|  | 13c | Describe any methods used to tabulate or visually display results of individual studies and syntheses. | Methods presentation in tables (Table 1, Table 2) and figures (Figure 2 forest plot). |
|  | 13d | Describe any methods used to synthesize results and provide a rationale for the choice(s). If meta-analysis was performed, describe the model(s), method(s) to identify the presence and extent of statistical heterogeneity, and software package(s) used. | Methods meta-analysis with fixed-effect model using R (meta, metafor packages). |
|  | 13e | Describe any methods used to explore possible causes of heterogeneity among study results (e.g. subgroup analysis, meta-regression). | Methods meta-regression for age and sex; sensitivity analysis. |
|  | 13f | Describe any sensitivity analyses conducted to assess robustness of the synthesized results. | Methods sensitivity analysis (leave-one-out). |
| Reporting bias assessment | 14 | Describe any methods used to assess risk of bias due to missing results in a synthesis (arising from reporting biases). | Methods publication bias assessment (funnel plot, Egger’s test). |
| Certainty assessment | 15 | Describe any methods used to assess certainty (or confidence) in the body of evidence for an outcome. | Not formally performed; certainty not graded (noted in Discussion Limitations). |
| **RESULTS** | | |  |
| Study selection | 16a | Describe the results of the search and selection process, from the number of records identified in the search to the number of studies included in the review, ideally using a flow diagram. | Results 'Study Selection'; includes PRISMA flow diagram (Figure 1). |
|  | 16b | Cite studies that might appear to meet the inclusion criteria, but which were excluded, and explain why they were excluded. | Results 'Study Selection'; excluded studies summarized (animal, unsuitable outcomes, etc.). |
| Study characteristics | 17 | Cite each included study and present its characteristics. | Results 'Study Characteristics' (Table 2). |
| Risk of bias in studies | 18 | Present assessments of risk of bias for each included study. | Results 'Risk of Bias' subsection (Table 1). |
| Results of individual studies | 19 | For all outcomes, present, for each study: (a) summary statistics for each group (where appropriate) and (b) an effect estimate and its precision (e.g. confidence/credible interval), ideally using structured tables or plots. | Results Tables 2–3 show individual study results (GCS, GOS, mortality). |
| Results of syntheses | 20a | For each synthesis, briefly summarise the characteristics and risk of bias among contributing studies. | Results 'Functional and Clinical Outcomes'; summary of included studies and risk of bias. |
|  | 20b | Present results of all statistical syntheses conducted. If meta-analysis was done, present for each the summary estimate and its precision (e.g. confidence/credible interval) and measures of statistical heterogeneity. If comparing groups, describe the direction of the effect. | Results 'Quantitative Synthesis (Meta-Analysis)' (SMD, CI, I²=0%). |
|  | 20c | Present results of all investigations of possible causes of heterogeneity among study results. | Results 'Meta-regression'; explores age and gender effects. |
|  | 20d | Present results of all sensitivity analyses conducted to assess the robustness of the synthesized results. | Results 'Sensitivity analysis'; confirmed result stability. |
| Reporting biases | 21 | Present assessments of risk of bias due to missing results (arising from reporting biases) for each synthesis assessed. | Results 'Publication bias'; funnel plot and Egger’s test, p=0.71. |
| Certainty of evidence | 22 | Present assessments of certainty (or confidence) in the body of evidence for each outcome assessed. | Not graded formally; discussed as moderate-quality evidence in Discussion. |
| **DISCUSSION** | | |  |
| Discussion | 23a | Provide a general interpretation of the results in the context of other evidence. | Discussion 'Summary of Main Findings' and 'Interpretation in Context of Previous Research'. |
|  | 23b | Discuss any limitations of the evidence included in the review. | Discussion 'Limitations of evidence'; small samples, heterogeneity. |
|  | 23c | Discuss any limitations of the review processes used. | Discussion 'Limitations of the review process'; incomplete blinding, small trials. |
|  | 23d | Discuss implications of the results for practice, policy, and future research. | Discussion 'Clinical Implications' and 'Future Directions'. |
| **OTHER INFORMATION** | | |  |
| Registration and protocol | 24a | Provide registration information for the review, including register name and registration number, or state that the review was not registered. | Methods 'Study Design and Registration'; PROSPERO ID: 1088575. |
|  | 24b | Indicate where the review protocol can be accessed, or state that a protocol was not prepared. | Methods protocol prepared a priori; not publicly posted. |
|  | 24c | Describe and explain any amendments to information provided at registration or in the protocol. | No amendments noted. |
| Support | 25 | Describe sources of financial or non-financial support for the review, and the role of the funders or sponsors in the review. | Declarations 'Funding'; no financial support received. |
| Competing interests | 26 | Declare any competing interests of review authors. | Declarations 'Competing interests'; none declared. |
| Availability of data, code and other materials | 27 | Report which of the following are publicly available and where they can be found: template data collection forms; data extracted from included studies; data used for all analyses; analytic code; any other materials used in the review. | Declarations ‘Availability of data and materials'; not applicable, data from published studies. |

*From:*  Page MJ, McKenzie JE, Bossuyt PM, Boutron I, Hoffmann TC, Mulrow CD, et al. The PRISMA 2020 statement: an updated guideline for reporting systematic reviews. BMJ 2021;372:n71. doi: 10.1136/bmj.n71. This work is licensed under CC BY 4.0. To view a copy of this license, visit <https://creativecommons.org/licenses/by/4.0/>
