## Supplementory for "Neuroprotective Effects of Vitamin D Supplementation on Outcomes in Traumatic Brain Injury: A Systematic Review and Meta-Analysis"

| **Study** | **Num of positive answers** | **Num of negative answers** | **Num of partial/unclear answers** | **Final status** |
| --- | --- | --- | --- | --- |
| **Randomized control trial studies** | | | | |
| Sharma et al. 2020 | 12 | - | 1 | Low |
| Razmkon et al. 2011 | 7 | 2 | 4 | Low |
| Arabi et al. 2020 | 5 | 4 | 2 | Moderate |
| Aminmansour et al. 2012 | 10 | 1 | 2 | Low |
| Zhang 2018 | 8 | - | 5 | Low |
| Masbough et al. 2024 | 8 | 1 | 4 | Low |
| Shafiei et al. 2022 | 11 | - | 2 | Low |
| **Cohort studies** | | | | |
| Lee et al. 2019 | 8 | - | 3 | Low |
| Guan et al. 2017 | 9 | 1 | 1 | Low |

Page 7:

*Table 1. The risk of bias assessment JBI Critical Appraisal checklists*
