## Supplementory for "Neuroprotective Effects of Vitamin D Supplementation on Outcomes in Traumatic Brain Injury: A Systematic Review and Meta-Analysis"

**Search results were extracted on January 16, 2025**

**Search strategies**

First search for all vitamins: 4546

Duplicate in EndNote: 960

First involved in Rayan: 3586

Duplication in Rayan: 600: not duplicated: 53 / resolved: 272 / deleted: 305

Final duplication: 1265

Primary screening (all vitamins): 3281

Vitamin E and Vitamin D: 234

- Exclude: 206
- Review: 32
- Case report: 7
- Animal study: 71
- Not related: 96

Second screening (full text): 28

- Include: 9
- Exclude:17
- Not found: 2

PubMed: 885

#1AND (#2 OR #3 OR #4) AND (#5 OR #6 OR #7 OR #8)

(((((((((((Treatment[MeSH Terms]) OR (Therapeutic[Title/Abstract])) OR (Therapy[Title/Abstract])) OR (Therapies[Title/Abstract])) OR (Treatments[Title/Abstract])) OR (Restoration[Title/Abstract])) OR (Improvement[Title/Abstract])) OR ((Management[MeSH Terms]) OR (Disease-Management[Title/Abstract]))) OR ((((Prognosis[MeSH Terms]) OR (Prognoses[Title/Abstract])) OR (Prognostic Factors[Title/Abstract])) OR (Prognostic Factor[Title/Abstract]))) OR (((Recovery of function[MeSH Terms]) OR (Function Recoveries[Title/Abstract])) OR (Function Recovery[Title/Abstract]))) AND (((((((((((Traumatic Brain injury[MeSH Terms]) OR (Traumatic Brain Injuries[Title/Abstract])) OR (Brain Trauma[Title/Abstract])) OR (Brain Traumas[Title/Abstract])) OR (Traumatic Brain Injury[Title/Abstract])) OR (Traumatic Encephalopathies[Title/Abstract])) OR (Traumatic Encephalopathy[Title/Abstract])) OR (TBIs[Title/Abstract])) OR (TBI[Title/Abstract])) OR ((((((((((((((((((((((((((((((Craniocerebral Trauma[MeSH Terms]) OR (Craniocerebral Traumas[Title/Abstract])) OR (Head Injuries[Title/Abstract])) OR (Head Injury[Title/Abstract])) OR (Craniocerebral Injuries[Title/Abstract])) OR (Craniocerebral Injury[Title/Abstract])) OR (Head Trauma[Title/Abstract])) OR (Head Traumas[Title/Abstract])) OR (Frontal Region Trauma[Title/Abstract])) OR (Frontal Region Traumas[Title/Abstract])) OR (Forehead Trauma[Title/Abstract])) OR (Forehead Traumas[Title/Abstract])) OR (Occipital Region Trauma[Title/Abstract])) OR (Occipital Region Traumas[Title/Abstract])) OR (Occipital Traum[Title/Abstract])) OR (Occipital Traumas[Title/Abstract])) OR (Parietal Region Trauma[Title/Abstract])) OR (Parietal Region Traumas[Title/Abstract])) OR (Temporal Region Trauma[Title/Abstract])) OR (Temporal Region Traumas[Title/Abstract])) OR (Crushing Skull Injury[Title/Abstract])) OR (Crushing Skull Injuries[Title/Abstract])) OR (Multiple Head Injury[Title/Abstract])) OR (Multiple Head Injuries[Title/Abstract])) OR (Minor Head Injuries[Title/Abstract])) OR (Minor Head Injury[Title/Abstract])) OR (Open Head Injuries[Title/Abstract])) OR (Open Head Injury[Title/Abstract])) OR (Superficial Head Injuries[Title/Abstract])) OR (Superficial Head Injury[Title/Abstract]))) OR ((((((((((((CONCUSSION[MeSH Terms]) OR (Brain Concussions[Title/Abstract])) OR (Cerebral Concussion[Title/Abstract])) OR (Cerebral Concussions[Title/Abstract])) OR (Commotio Cerebri[Title/Abstract])) OR (Intermediate Concussion[Title/Abstract])) OR (Intermediate Concussions[Title/Abstract])) OR (Mild Concussion[Title/Abstract])) OR (Mild Concussions[Title/Abstract])) OR (Mild Traumatic Brain Injury[Title/Abstract])) OR (Severe Concussion[Title/Abstract])) OR (Severe Concussions[Title/Abstract])))) AND (((((((((((((((((((((((((((((((((((((((((((((((((((((((((((((((((((((((((((((((Vitamin[MeSH Terms]) OR (choline[Title/Abstract])) OR (dehydrocholic acid[Title/Abstract])) OR (magnesium orotate[Title/Abstract])) OR (orotic acid[Title/Abstract])) OR (Vitamin B12 drug combination[Title/Abstract])) OR (coenzyme Q10[Title/Abstract])) OR (vitamin A[Title/Abstract])) OR (pentifyllin[Title/Abstract])) OR (nicotinic acid[Title/Abstract])) OR (vitamin E combination[Title/Abstract])) OR (aminopyrine[Title/Abstract])) OR (dexamethasone[Title/Abstract])) OR (phenylbutazone[Title/Abstract])) OR (thiamine[Title/Abstract])) OR (1 alpha-hydroxyergocalciferol[Title/Abstract])) OR (calcium ascorbate[Title/Abstract])) OR (Tocovid[Title/Abstract])) OR (Ascorbic Acid[Title/Abstract])) OR (Calcifediol[Title/Abstract])) OR (Calcitriol[Title/Abstract])) OR (Cholecalciferol[Title/Abstract])) OR (Cod Liver Oil[Title/Abstract])) OR (Dehydroascorbic Acid[Title/Abstract])) OR (Dihydrotachysterol[Title/Abstract])) OR (Dihydroxycholecalciferols[Title/Abstract])) OR (Ergocalciferols[Title/Abstract])) OR (Hydroxycholecalciferols[Title/Abstract])) OR (Vitamin K-1[Title/Abstract])) OR (Vitamin D[Title/Abstract])) OR (Vitamin E[Title/Abstract])) OR (Vitamin K[Title/Abstract])) OR (Vitamin U[Title/Abstract])) OR (24,25-Dihydroxyvitamin D 3[Title/Abstract])) OR (25-Hydroxyvitamin D 2[Title/Abstract])) OR (Vitamin K-2[Title/Abstract])) OR (Vitamin K-3[Title/Abstract])) OR (alpha-Tocopherol[Title/Abstract])) OR (beta-Tocopherol[Title/Abstract])) OR (gamma-Tocopherol[Title/Abstract])) OR (Tocopherols[Title/Abstract])) OR (Tocotrienols[Title/Abstract])) OR (provitamin C[Title/Abstract])) OR (7-dehydrocholesterol[Title/Abstract])) OR (sapotexanthin[Title/Abstract])) OR (alpha-cryptoxanthin[Title/Abstract])) OR (Beta-Cryptoxanthin[Title/Abstract])) OR (Dehydrocholesterols[Title/Abstract])) OR (beta Carotene[Title/Abstract])) OR (Ergosterol[Title/Abstract])) OR (Palmitoylcarnitine[Title/Abstract])) OR (Pantothenic Acid[Title/Abstract])) OR (Tetrahydrofolates[Title/Abstract])) OR (Thiamine[Title/Abstract])) OR (Thiamine Monophosphate[Title/Abstract])) OR (Thiamine Pyrophosphate[Title/Abstract])) OR (Thiamine Triphosphate[Title/Abstract])) OR (Thioctic Acid[Title/Abstract])) OR (Pteroylpolyglutamic Acids[Title/Abstract])) OR (Pyridoxal[Title/Abstract])) OR (Pyridoxal Phosphate[Title/Abstract])) OR (Pyridoxamine[Title/Abstract])) OR (Pyridoxine[Title/Abstract])) OR (Niacin[Title/Abstract])) OR (Niacinamide[Title/Abstract])) OR (Biotin[Title/Abstract])) OR (Leucovorin[Title/Abstract])) OR (Cobamides[Title/Abstract])) OR (Nicorandil[Title/Abstract])) OR (Flavin Mononucleotide[Title/Abstract])) OR (Folic Acid[Title/Abstract])) OR (Formyltetrahydrofolates[Title/Abstract])) OR (Fursultiamin[Title/Abstract])) OR (Riboflavin[Title/Abstract])) OR (Hydroxocobalamin[Title/Abstract])) OR (Vitamin B 12[Title/Abstract])) OR (Vitamin B 6[Title/Abstract])) OR (Inositol[Title/Abstract])) OR (Acetylcarnitine[Title/Abstract]))

Scopus: 2353

(”Vitamin“ OR “choline” OR “dehydrocholic acid” OR “magnesium orotate” OR “orotic acid” OR “vitamin B12 drug combination” OR “coenzyme Q10” OR “vitamin A” OR “pentifyllin” OR “nicotinic acid” OR “vitamin E combination” OR “aminopyrine” OR “dexamethasone” OR “phenylbutazone” OR “thiamine” OR “1 alpha-hydroxyergocalciferol” OR “ calcium ascorbate” OR “tocovid” OR “ascorbic acid” OR “calcifediol” OR “calcitriol” OR “cholecalciferol” OR “cod liver oil” OR “dehydroascorbic acid” OR “dihydrotachysterol” OR “dihydroxycholecalcalciferols” OR “ergocalciferols” OR “hydroxycholecalciferols” OR “vitamin K 1” OR “ vitamin D” OR “vitamin E” OR “vitamin K” OR “vitamin U” OR “24,25-Dihydroxyvitamin D 3” OR “25-Hydroxyvitamin D 2” OR “Vitamin K 2” OR “Vitamin K 3” OR “alpha-Tocopherol” OR “beta-Tocopherol” OR “gamma-Tocopherol” OR “Tocopherols” OR “Tocotrienols” OR “provitamin C” OR “7-dehydrocholesterol” OR “sapotexanthin” OR “alpha-cryptoxanthin” OR “Beta-Cryptoxanthin” OR “Dehydrocholesterols” OR “beta Carotene” OR “Ergosterol” OR “Palmitoylcarnitine” OR “Pantothenic Acid” OR “Tetrahydrofolates” OR “Thiamine” OR “Thiamine Monophosphate” OR “Thiamine Pyrophosphate” OR “Thiamine Triphosphate” OR “Thioctic Acid” OR “Pteroylpolyglutamic Acids” OR “Pyridoxal” OR “Pyridoxal Phosphate” OR “Pyridoxamine” OR “Pyridoxine” OR “Niacin” OR “Niacinamide” OR “Biotin” OR “Leucovorin” OR “Cobamides” OR “Nicorandil” OR “Flavin Mononucleotide” OR “Folic Acid” OR “Formyltetrahydrofolates” OR “Fursultiamin” OR “Riboflavin” OR “Hydroxocobalamin” OR “Vitamin B 12” OR “Vitamin B 6” OR “Inositol” OR “Acetylcarnitine”) AND ((“Traumatic Brain injury” OR “Traumatic Brain Injuries” OR “Brain Trauma” OR “Brain Traumas” OR “Traumatic Brain Injury” OR “Traumatic Encephalopathies” OR “Traumatic Encephalopathy” OR “TBIs” OR “TBI”) OR (“Craniocerebral Trauma” OR “Craniocerebral Traumas” OR “Head Injuries” OR “Head Injury” OR “Craniocerebral Injuries” OR “Craniocerebral Injury” OR “Head Trauma” OR “Head Traumas” OR “Frontal Region Trauma” OR “Frontal Region Traumas” OR “Forehead Trauma” OR “Forehead Traumas” OR “Occipital Region Trauma” OR “Occipital Region Traumas” OR “Occipital Trauma” OR “Occipital Traumas” OR “Parietal Region Trauma” OR “Parietal Region Traumas” OR “Temporal Region Trauma” OR “Temporal Region Traumas” OR “Crushing Skull Injury” OR “Crushing Skull Injuries” OR “Multiple Head Injury” OR “Multiple Head Injuries” OR “Minor Head Injuries” OR “Minor Head Injury” OR “Open Head Injuries” OR “Open Head Injury” OR “Superficial Head Injuries” OR “Superficial Head Injury”) OR (“CONCUSSION” OR “Brain Concussions” OR “Concussion, Brain” OR “Cerebral Concussion” OR “Cerebral Concussions” OR “Concussion, Cerebral” OR “Commotio Cerebri” OR “Concussion, Intermediate” OR “Intermediate Concussion” OR “Intermediate Concussions” OR “Concussion, Mild” OR “Mild Concussion” OR “Mild Concussions” OR “Mild Traumatic Brain Injury” OR “Concussion, Severe” OR “Severe Concussion” OR “Severe Concussions”)) AND ((“Treatment” OR “Therapeutic” OR “Therapy” OR “Therapies” OR “Treatment” OR “Treatments” OR “Restoration” OR “Improvement”) OR (“Management” OR “Disease Managements”) OR ("Prognosis” OR “Prognoses” OR “Prognostic Factors” OR “Prognostic Factor”) OR (“Recovery of function” OR “Function Recoveries” OR “Function Recovery”))

Embase: 591

(('vitamin':ti,ab OR 'choline':ti,ab OR 'dehydrocholic acid':ti,ab OR 'magnesium orotate':ti,ab OR 'orotic acid':ti,ab OR 'vitamin B12 drug combination':ti,ab OR 'coenzyme Q10':ti,ab OR 'vitamin A':ti,ab OR 'pentifyllin':ti,ab OR 'nicotinic acid':ti,ab OR 'vitamin E combination':ti,ab OR 'aminopyrine':ti,ab OR 'dexamethasone':ti,ab OR 'phenylbutazone':ti,ab OR 'thiamine':ti,ab OR '1 alpha-hydroxyergocalciferol':ti,ab OR 'calcium ascorbate':ti,ab OR 'tocovid':ti,ab OR 'ascorbic acid':ti,ab OR 'calcifediol':ti,ab OR 'calcitriol':ti,ab OR 'cholecalciferol':ti,ab OR 'cod liver oil':ti,ab OR 'dehydroascorbic acid':ti,ab OR 'dihydrotachysterol':ti,ab OR 'dihydroxycholecalcalciferols':ti,ab OR 'ergocalciferols':ti,ab OR 'hydroxycholecalciferols':ti,ab OR 'vitamin K1':ti,ab OR 'vitamin D':ti,ab OR 'vitamin E':ti,ab OR 'vitamin K':ti,ab OR 'vitamin U':ti,ab OR '24,25-dihydroxyvitamin D3':ti,ab OR '25-hydroxyvitamin D2':ti,ab OR 'vitamin K2':ti,ab OR 'vitamin K3':ti,ab OR 'alpha-tocopherol':ti,ab OR 'beta-tocopherol':ti,ab OR 'gamma-tocopherol':ti,ab OR 'tocopherols':ti,ab OR 'tocotrienols':ti,ab OR 'provitamin C':ti,ab OR '7-dehydrocholesterol':ti,ab OR 'sapotexanthin':ti,ab OR 'alpha-cryptoxanthin':ti,ab OR 'beta-cryptoxanthin':ti,ab OR 'dehydrocholesterols':ti,ab OR 'beta carotene':ti,ab OR 'ergosterol':ti,ab OR 'palmitoylcarnitine':ti,ab OR 'pantothenic acid':ti,ab OR 'tetrahydrofolates':ti,ab OR 'thiamine monophosphate':ti,ab OR 'thiamine pyrophosphate':ti,ab OR 'thiamine triphosphate':ti,ab OR 'thioctic acid':ti,ab OR 'pteroylpolyglutamic acids':ti,ab OR 'pyridoxal':ti,ab OR 'pyridoxal phosphate':ti,ab OR 'pyridoxamine':ti,ab OR 'pyridoxine':ti,ab OR 'niacin':ti,ab OR 'niacinamide':ti,ab OR 'biotin':ti,ab OR 'leucovorin':ti,ab OR 'cobamides':ti,ab OR 'nicorandil':ti,ab OR 'flavin mononucleotide':ti,ab OR 'folic acid':ti,ab OR 'formyltetrahydrofolates':ti,ab OR 'fursultiamin':ti,ab OR 'riboflavin':ti,ab OR 'hydroxocobalamin':ti,ab OR 'vitamin B12':ti,ab OR 'vitamin B6':ti,ab OR 'inositol':ti,ab OR 'acetylcarnitine':ti,ab) AND ('traumatic brain injury':ti,ab OR 'brain trauma':ti,ab OR 'TBIs':ti,ab OR 'concussion':ti,ab OR 'head injury':ti,ab OR 'craniocerebral trauma':ti,ab) AND ('treatment':ti,ab OR 'therapy':ti,ab OR 'management':ti,ab OR 'recovery':ti,ab OR 'prognosis':ti,ab))

Web of science: 717

(”Vitamin“ OR “choline” OR “dehydrocholic acid” OR “magnesium orotate” OR “orotic acid” OR “vitamin B12 drug combination” OR “coenzyme Q10” OR “vitamin A” OR “pentifyllin” OR “nicotinic acid” OR “vitamin E combination” OR “aminopyrine” OR “dexamethasone” OR “phenylbutazone” OR “thiamine” OR “1 alpha-hydroxyergocalciferol” OR “ calcium ascorbate” OR “tocovid” OR “ascorbic acid” OR “calcifediol” OR “calcitriol” OR “cholecalciferol” OR “cod liver oil” OR “dehydroascorbic acid” OR “dihydrotachysterol” OR “dihydroxycholecalcalciferols” OR “ergocalciferols” OR “hydroxycholecalciferols” OR “vitamin K 1” OR “ vitamin D” OR “vitamin E” OR “vitamin K” OR “vitamin U” OR “24,25-Dihydroxyvitamin D 3” OR “25-Hydroxyvitamin D 2” OR “Vitamin K 2” OR “Vitamin K 3” OR “alpha-Tocopherol” OR “beta-Tocopherol” OR “gamma-Tocopherol” OR “Tocopherols” OR “Tocotrienols” OR “provitamin C” OR “7-dehydrocholesterol” OR “sapotexanthin” OR “alpha-cryptoxanthin” OR “Beta-Cryptoxanthin” OR “Dehydrocholesterols” OR “beta Carotene” OR “Ergosterol” OR “Palmitoylcarnitine” OR “Pantothenic Acid” OR “Tetrahydrofolates” OR “Thiamine” OR “Thiamine Monophosphate” OR “Thiamine Pyrophosphate” OR “Thiamine Triphosphate” OR “Thioctic Acid” OR “Pteroylpolyglutamic Acids” OR “Pyridoxal” OR “Pyridoxal Phosphate” OR “Pyridoxamine” OR “Pyridoxine” OR “Niacin” OR “Niacinamide” OR “Biotin” OR “Leucovorin” OR “Cobamides” OR “Nicorandil” OR “Flavin Mononucleotide” OR “Folic Acid” OR “Formyltetrahydrofolates” OR “Fursultiamin” OR “Riboflavin” OR “Hydroxocobalamin” OR “Vitamin B 12” OR “Vitamin B 6” OR “Inositol” OR “Acetylcarnitine”) **AND** ((“Traumatic Brain injury” OR “Traumatic Brain Injuries” OR “Brain Trauma” OR “Brain Traumas” OR “Traumatic Brain Injury” OR “Traumatic Encephalopathies” OR “Traumatic Encephalopathy” OR “TBIs” OR “TBI”) **OR** (“Craniocerebral Trauma” OR “Craniocerebral Traumas” OR “Head Injuries” OR “Head Injury” OR “Craniocerebral Injuries” OR “Craniocerebral Injury” OR “Head Trauma” OR “Head Traumas” OR “Frontal Region Trauma” OR “Frontal Region Traumas” OR “Forehead Trauma” OR “Forehead Traumas” OR “Occipital Region Trauma” OR “Occipital Region Traumas” OR “Occipital Trauma” OR “Occipital Traumas” OR “Parietal Region Trauma” OR “Parietal Region Traumas” OR “Temporal Region Trauma” OR “Temporal Region Traumas” OR “Crushing Skull Injury” OR “Crushing Skull Injuries” OR “Multiple Head Injury” OR “Multiple Head Injuries” OR “Minor Head Injuries” OR “Minor Head Injury” OR “Open Head Injuries” OR “Open Head Injury” OR “Superficial Head Injuries” OR “Superficial Head Injury”) **OR** (“CONCUSSION” OR “Brain Concussions” OR “Concussion, Brain” OR “Cerebral Concussion” OR “Cerebral Concussions” OR “Concussion, Cerebral” OR “Commotio Cerebri” OR “Concussion, Intermediate” OR “Intermediate Concussion” OR “Intermediate Concussions” OR “Concussion, Mild” OR “Mild Concussion” OR “Mild Concussions” OR “Mild Traumatic Brain Injury” OR “Concussion, Severe” OR “Severe Concussion” OR “Severe Concussions”)) **AND** ((“Treatment” OR “Therapeutic” OR “Therapy” OR “Therapies” OR “Treatment” OR “Treatments” OR “Restoration” OR “Improvement”) **OR** (“Management” OR “Disease Managements”) **OR** ("Prognosis” OR “Prognoses” OR “Prognostic Factors” OR “Prognostic Factor”) **OR** (“Recovery of function” OR “Function Recoveries” OR “Function Recovery”))

**Keywords:**

**#1 Vitamin: During the first screening, all keywords were preserved. After duplicate entries were removed, the YELLOW-highlighted words were selected.**

choline,

dehydrocholic acid,

magnesium orotate,

orotic acid,

Vitamin B12 drug combination

coenzyme Q10

vitamin A,

pentifyllin,

nicotinic acid,

vitamin E combination

aminopyrine,

dexamethasone,

phenylbutazone,

thiamine,

1 alpha-hydroxyergocalciferol

calcium ascorbate

Tocovid

Ascorbic Acid

Calcifediol

Calcitriol

Cholecalciferol

Cod Liver Oil

Dehydroascorbic Acid

Dihydrotachysterol

Dihydroxycholecalciferols

Ergocalciferols

Hydroxycholecalciferols

Vitamin K 1

Vitamin D

Vitamin E

Vitamin K

Vitamin U

24,25-Dihydroxyvitamin D 3

25-Hydroxyvitamin D 2

Vitamin K 2

Vitamin K 3

alpha-Tocopherol

beta-Tocopherol

gamma-Tocopherol

Tocopherols

Tocotrienols

provitamin C

7-dehydrocholesterol

sapotexanthin

alpha-cryptoxanthin

Beta-Cryptoxanthin

Dehydrocholesterols

beta Carotene

Ergosterol

Palmitoylcarnitine

Pantothenic Acid

Tetrahydrofolates

Thiamine

Thiamine Monophosphate

Thiamine Pyrophosphate

Thiamine Triphosphate

Thioctic Acid

Pteroylpolyglutamic Acids

Pyridoxal

Pyridoxal Phosphate

Pyridoxamine

Pyridoxine

Niacin

Niacinamide

Biotin

Leucovorin

Cobamides

Nicorandil

Flavin Mononucleotide

Folic Acid

Formyltetrahydrofolates

Fursultiamin

Riboflavin

Hydroxocobalamin

Vitamin B 12

Vitamin B 6

Inositol

Acetylcarnitine

**#2 Traumatic Brain injury:**

- Traumatic Brain Injuries
- Brain Trauma
- Brain Traumas
- Traumatic Brain Injury
- Traumatic Encephalopathies
- Traumatic Encephalopathy
- TBIs
- TBI

**#3 Craniocerebral Trauma:**

- Craniocerebral Traumas
- Head Injuries
- Head Injury
- Craniocerebral Injuries
- Craniocerebral Injury
- Head Trauma
- Head Traumas
- Frontal Region Trauma
- Frontal Region Traumas
- Forehead Trauma
- Forehead Traumas
- Occipital Region Trauma
- Occipital Region Traumas
- Occipital Trauma
- Occipital Traumas
- Parietal Region Trauma
- Parietal Region Traumas
- Temporal Region Trauma
- Temporal Region Traumas
- Crushing Skull Injury
- Crushing Skull Injuries
- Multiple Head Injury
- Multiple Head Injuries
- Minor Head Injuries
- Minor Head Injury
- Open Head Injuries
- Open Head Injury
- Superficial Head Injuries
- Superficial Head Injury

**#4 CONCUSSION:**

- Brain Concussions
- Concussion, Brain
- Cerebral Concussion
- Cerebral Concussions
- Concussion, Cerebral
- Commotio Cerebri
- Concussion, Intermediate
- Intermediate Concussion
- Intermediate Concussions
- Concussion, Mild
- Mild Concussion
- Mild Concussions
- Mild Traumatic Brain Injury
- Concussion, Severe
- Severe Concussion
- Severe Concussions

**#5 Treatment:**

- Therapeutic
- Therapy
- Therapies
- Treatment
- Treatments
- Restoration
- Improvement

**#6 Management:**

- Disease Managements

**#7 Prognosis:**

- Prognoses
- Prognostic Factors
- Prognostic Factor

**#8 Recovery of function**:

- Function Recoveries
- Function Recovery
