## Supplementory for "Neuroprotective Effects of Vitamin D Supplementation on Outcomes in Traumatic Brain Injury: A Systematic Review and Meta-Analysis"

***Table 3*** *Meta-regression results for the influence of age and gender on GCS outcomes*

| Covariate | No. of Studies | Estimate | Estimate SE | P Value | R² | τ² | I² |
| --- | --- | --- | --- | --- | --- | --- | --- |
| Age | 3 | 0.021 | 0.0544 | 0.701 | 0 | 0.094 | 44.258 |
| Female | 3 | 16.319 | 12.125 | 0.178 | 100 | 0 | 0 |
